# Publish or perish: reporting characteristics of peer-reviewed publications, pre-prints and registered studies on the COVID-19 pandemic

**DOI:** 10.1101/2020.06.14.20130823

**Authors:** S Gianola, TS Jesus, S Bargeri, G Castellini

## Abstract

**Background:** The COVID-19 pandemic has resulted in a mass of academic papers being published in a very brief span of time. Our aim was to compare the amount and reporting characteristics of COVID-19 related peer-reviewed and pre-prints publications. We also investigated the amount of ongoing trials and systematic reviews.

**Methods and findings:** A cross-sectional study of publications covering the COVID-19 pandemic time frame, up to May 20, 2020 was conducted. PubMed with appropriate combinations of Medical Subject Headings and COVID-19 section of MedRxiv and BioRxiv archives were searched. We examined Clinicaltrial.gov, Chinese Clinical Trial Registry, EU Register and 15 other trial registers as well as the international prospective register of systematic reviews (PROSPERO). Characteristics of each publication source were extracted. Regression analyses and Z tests were used to analyze publication trends over the weeks and compare their relative proportions.

We found 3635 peer-reviewed publications and 3805 pre-prints, of which 8.6% (n=329) were published in indexed journals. Peer-reviewed and pre-print publications amount both increased significantly over time (p<0.001). Case reports (peer-reviewed: 6% vs pre-prints: 0.9%, *p*<0.001) and letters (17.4% vs 0.5%, *p*<0.001) accounted for a greater share of the peer-reviewed compared to pre-print publications. In turn, randomized controlled trials (0.22% vs 0.63% *p*<0.001) and systematic reviews (0.08% vs 5%) accounted for a significantly greater share of the pre-print publications. Clinicaltrials.gov, Chinese Clinical Trial Registry and EU register included 57.9%, 49.5 % and 98.9% trials mostly still “recruiting”. PROSPERO amounted to 962 systematic review protocols.

**Conclusion:** Pre-prints were slightly more prevalent than peer-reviewed publications, yet both are growing. To fill the void given by the absence of published primary studies, immediate opinions (i.e., letters) has virulently been published in PubMed. However, preprints has been promoted as rapid responses to give direct and promptly access at scientific findings in this pandemic.

## Introduction

A novel human coronavirus named by the World Health Organization as Severe Acute Respiratory Syndrome coronavirus 2 (SARS-COV-2) has been causing a pandemic of flu-like disease from the end of 2019 (1). The global health emergency asked for urgent research priorities needed to inform the public health response to the worldwide spread of 2019-nCoV infections (2). However, the enormity of the scientific efforts and the speed at which the knowledge on this topic has been generated pose significant difficulties for everyone to stay abreast of these developments (3). SARS-COV-2 has sparked a parallel viral spread: science is being conducted, posted, and shared at an unprecedented rate. A bibliometric and scientometric analysis of publications can quantitatively examine the research progress of any topic and offer a comprehensive assessment of scientific research trends, which is widely used for mapping knowledge in different scientific disciplines (4, 5). In an effort to address challenges in Coronavirus Diseases 2019 (COVID-19) era and to better organize the emerging and rapidly scientific developments, scoping reviews with a bibliometric and scientometric analyses have been conducted (3, 6, 7) showing and visualizing the networks of contributing authors, institutions, and countries. However, none of these reviews have focused on epidemiology and reporting characteristics (8, 9) of this global burden assessing the evolution of research over the pandemic time (10-12). In the COVID-19 burden, the scientific effort has been spread in different sources: registers of primary studies (i.e, trials) and systematic reviews, preprints and peer-reviewed publications.

A clinical trials registry is a platform which catalogs registered clinical trials. Clinical trials can play an important role in the COVID-19 to translate evidence of experimental and observational researches into clinical practice (13, 14). Whereas, a systematic review protocol registry is an international database which prospectively lists systematic reviews in health and social care. Systematic reviews should be registered at inception (i.e. at the protocol stage) to help avoid outcome reporting biases, publication bias, unplanned duplication and waste of resource, especially true in a period of global health emergency (15, 16). Preprints are “preliminary reports of work that have not been certified by peer review, at least yet. Hence, they should not be relied on to guide clinical practice or health-related behavior and should not be reported in news media as established information” (17). Preprint servers are open access online repositories housing preprint research articles that on one hand enable authors to make their research immediately and freely available to receive commentary and peer review prior to journal submission accelerating the dissemination of scientific findings (18). Preprints posted during the Ebola and Zika outbreaks included novel analyses and new data, and most of those that were matched to peer-reviewed publications were available more than 100 days before publication (18). On the other hand, many pre-prints never become certified by peer review: indeed, less than 5% of Ebola and Zika journal articles were posted as preprints prior to publication in journals (18). Therefore, information in preprints lack the scrutiny and validity of an external, scientific review (19). In the pandemic era, an analysis of preprints and ongoing researches (i.e., trials and reviews) can be important since timeliness is key, even though peer reviews are expedited for new relevant research.

### Aims

In the context above, we aim to provide a snapshot of the amount of scientific development on COVID-19 worldwide, which may inform the current status of global research and provide insights on the needs for future research in terms of its amount, design, publication venues, and reporting characteristics. More specifically, we aim to do so by:

1. Comparing the amount of COVID-19 related research in peer-reviewed publications or pre-prints (not peer-reviewed), including as stratified by key reported characteristics: species (ie. humans, animals), study design (such as systematic reviews (SRs), randomized controlled trials (RCTs)) and research area (such as drug treatment).
2. Inspect the amount and key reported characteristics of COVID-19 ongoing research in the registers of clinical trials and systematic reviews (e.g., research Area).

### Methods

Design: Cross-sectional study of publications and its key study and reporting characteristics COVID-19 related publications until May 20 2020.

### Data sources

We searched for COVID-19 peer-reviewed publications in PubMed database through its indexation system. PubMed is a comprehensive research database comprised of more than 30 million citations, as of May 2020, for biomedical literature from MEDLINE, life science journals, and online books (20). In the health field, there is evidence that adding databases other than PubMed only had modest impact on SR results (21). In PubMed, Medical Subject Headings (MeSH) are organized in a hierarchical tree and assigned to each paper by subject-specialist indexers. For COVID-19, we used the Supplementary Concept Records (SCRs). MEDLINE indexers regularly come across substances in the literature that are not currently MeSH headings. When this happens, the National Library of Medicine staff adds these substances to the MeSH vocabulary as Supplementary Concept Records (SCRs). While MeSH headings are updated annually, new SCRs are added weekly. Therefore, COVID-19 articles are systemically indexed by research topic regardless of the specific words used by the authors. We conducted target searches in PubMed that included the use appropriated combinations of MeSH terms and SCRs (i.e. search filters) related to COVID-19 and related terms (e.g. SARS-COV-2, coronavirus disease), filtered for specie, research designs, and research area (See **S1 Appendix. PubMed database**).

We searched the two popular sources for COVID-19 related preprints, MedRxiv and BioRxiv databases, proxy indicators of scientific not peer-reviewed literature. Papers are examined by in-house staff who check for issues such as plagiarism and incompleteness (22) (**see S1 Appendix. Preprints MedRxiv and BioRxiv database**).

We then investigated all primary registries that meet the requirements of the International Committee of Medical Journal Editors (ICMJE) according to the WHO Registry Network. Primary registries in the WHO Registry Network meet specific criteria for content, quality and validity, accessibility, unique identification, technical capacity and administration (23). Additionally, we searched for International prospective register of systematic reviews focusing on PROSPERO database (**see S1 Appendix. Primary register; S1 Appendix. PROSPERO register**).

The whole searches in all sources were run in May 20, 2020 without restrictions on languages.

### Data extraction

Data records obtained from PubMed, MedRxiv and BioRxiv, Trials registers and PROSPERO database were imported to Excel for further grouping and analyses. Then the whole data extraction was performed by one author and checked by the second author. Any uncertainties were discussed among the data extractors.

For the peer-reviewed publications and pre-prints, we extracted the bibliometric data and then analyzed and summarized the subsets focusing on the following study or reporting characteristics: specie (i.e., humans, animals), publication type (i.e., SRs, RCTs, Epidemiologic Studies, Letter), the distribution by research area such as vaccine, treatment drug treatment, rehabilitation, diagnostic testing, measures for infection prevention and control (i.e., social distance, masking), biology, according to the indexation facilities and the respective search tags or filters. Since in the pre-prints no such filters/tags exists, we manually extracted and coded the above topics. The study designs such as laboratory experiments (e.g., genomics), prognostic models were included in the epidemiology studies.

For the primary registers (e.g., trials), the following characteristics were filtered/tagged and extracted: country, study type, phase, recruiting status. For data of PROSPERO database species and research area/tag have been extracted.

### Data analysis

Initially, descriptive statistics were used for the analysis. Over the volume of publications for each data element (e.g. publication type), percentages and frequencies were computed and displayed in tabular formats, paired between peer-reviewed and pre-print sources. After that, the amount of publications over time (weekly) was plotted into a run chart. Then, simple linear regressions methods with the ANOVA were used to analyze the growth of the number of COVID-19 publications over the weeks. Finally, two-sample Z-tests, two tails, analyzed whether the proportion of each characteristics of peer-reviewed source (i.e., PubMed) significantly differed from those of pre-print sources. P values <0.05 were considered significant. The data were stored and analyzed using STATA 16 software.

## Results

We respond to each of our two specific study objectives in the respective subsections below.

### 1) Amount and Characteristics of COVID-19 research peer vs not peer-reviewed (pre-print) publications

From December 2019 to May 2020, a total of 7440 COVID-19 related publications were found. From these, 3635 (48.9%) were peer-reviewed, and 3805 (51.1%) were preprints. **Fig 1** reports the weekly amount of both peer-reviewed and pre-print publications, from December 2019 to May 2020. The increase of peer-reviewed and pre-print publications are both statistically significant over time (peer-reviewed publications: *p<*0.001 r^2^=0.8239, and pre-prints: *p*<0.001 r^2^=0.9133, respectively). **Figs 2 and 3** shows the absolute and relative frequencies of peer-reviewed and pre-prints publications over each week. **Fig 4** shows the trend of the ratio between relative frequencies of peer-reviewed and pre-prints publications over weeks. The difference of the increase rate among relative frequencies between the two groups was found not statistically significant over weeks (*p*=0.2388, r^2^=0.0856).

**Figure 1.**
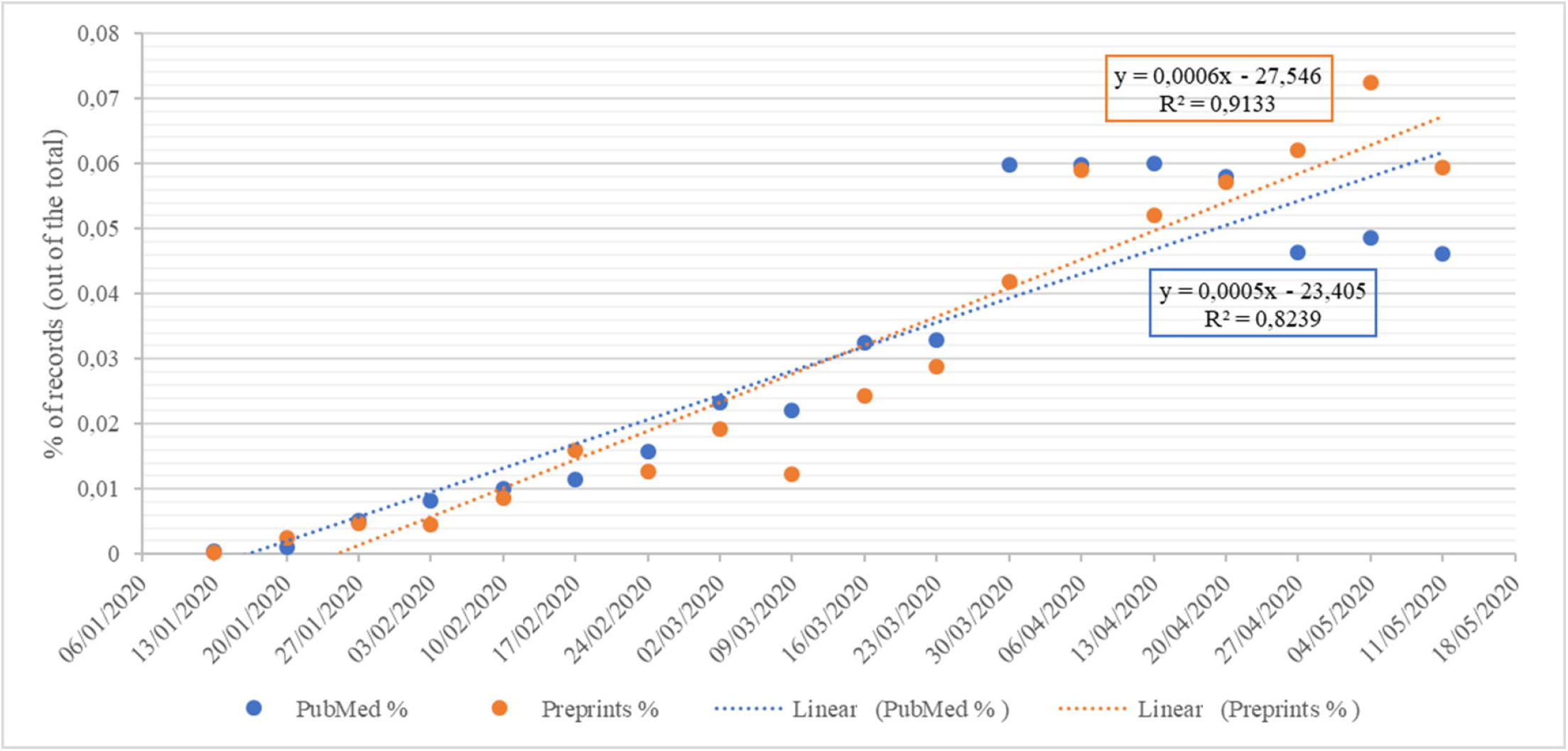
Linear regression over time for peer-reviewed and pre-print publications.

**Figure 2.**
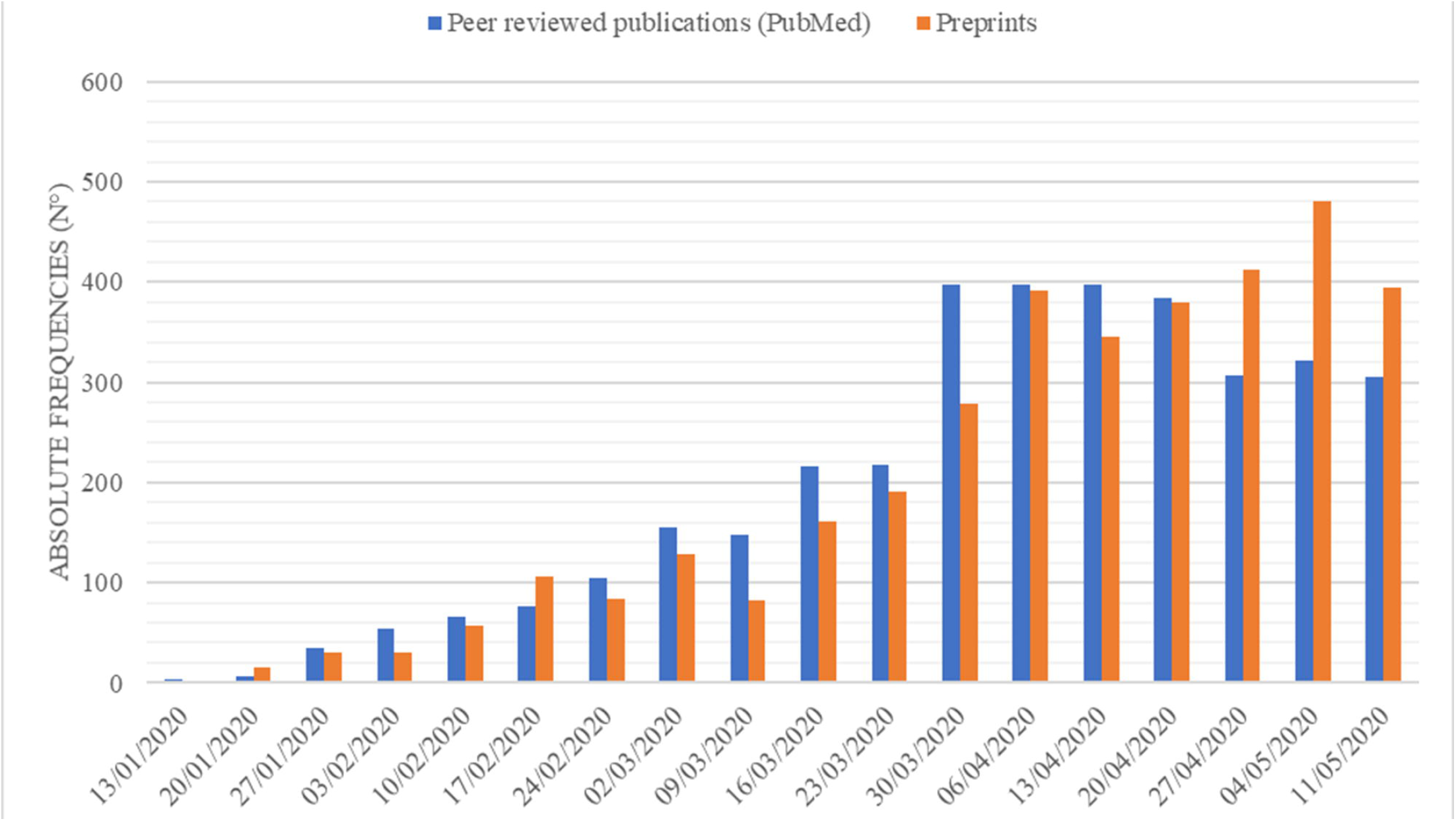
Absolute frequencies of peer-reviewed and pre-print publications over weeks.

**Figure 3.**
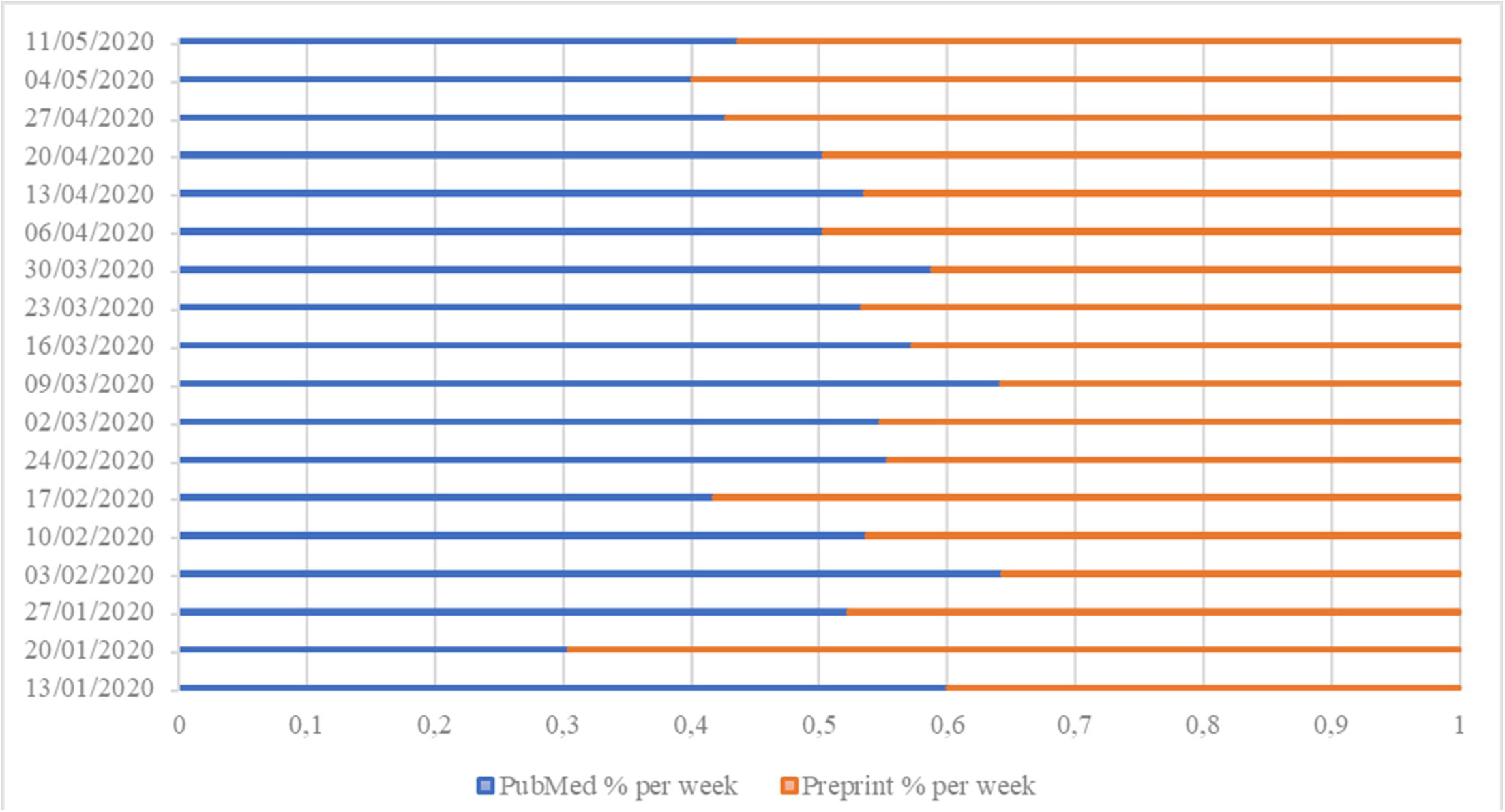
Relative distribution of peer-reviewed and pre-print publications per week.

**Figure 4.**
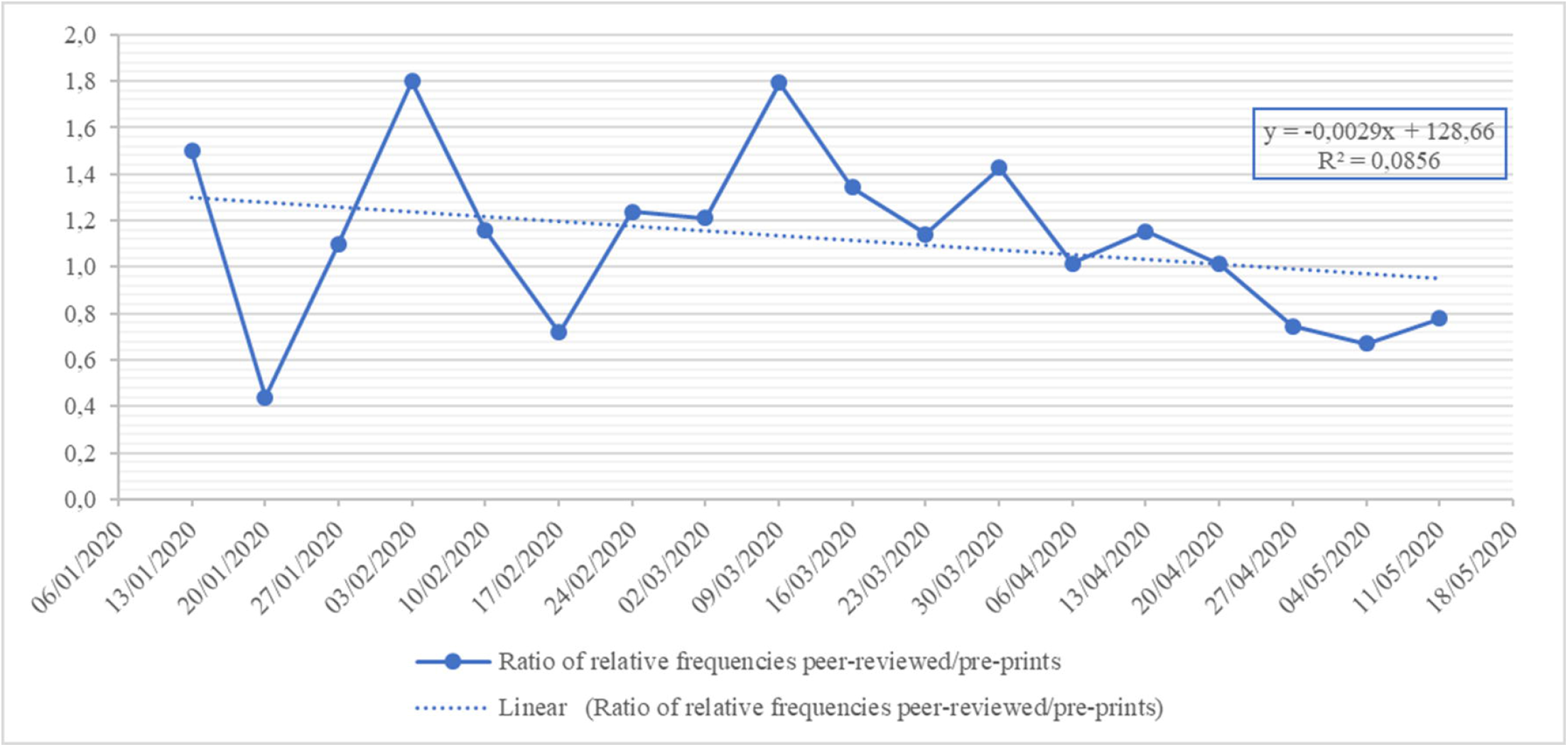
Linear regression of ratios between relative frequencies (peer-reviewed/pre-print publications) over weeks.

**Table 1** shows the reporting characteristics for both peer-reviewed and pre-prints COVID-19 related publications. In both peer-reviewed publications and pre-prints, “human” subjects were predominant as well as the most addressed research areas were prevention and control (26.1%, 950/3635 peer-reviewed vs 42.4%, 1615/3805 pre-prints; *p*<0.001) and diagnosis (21.5%, 781/3635 peer-reviewed vs 25.3%, 962/3805 pre-prints; *p*<0.001).

**Table 1.**
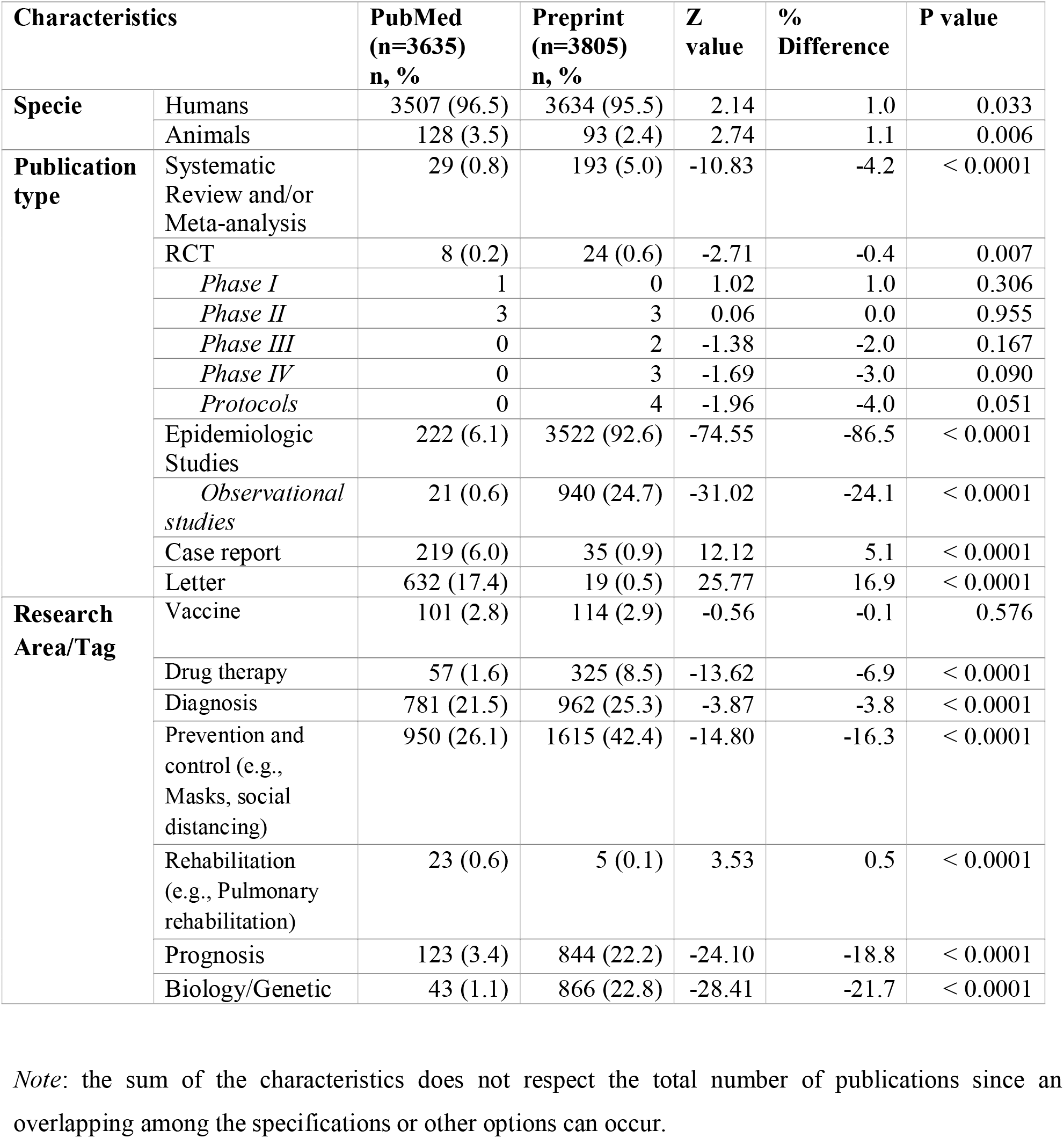
Characteristics of COVID-19 peer-reviewed (PubMed) vs not peer-reviewed (Preprints) publications.

Regarding the relative prevalence of study types, statistically significant differences were found among peer-reviewed and pre-print publications. For instance, RCT designs were significantly more common among pre-prints than among the peer-reviewed research (peer-reviewed: 0.2%; 8/3635 vs pre-prints: 0.6%, 24/3805; P<0.001). Similarly, SRs were significantly more common among preprints (peer-reviewed: 0.8%, 29/3635 vs pre-prints: 5.0%, 193/3805; P<0.001). Observational studies were also significantly more common in pre-print research than peer-reviewed publications (peer-reviewed: 0.6%, 21/3635 vs pre-prints: 24.7%, 940/3805; P<0.001). For case reports and letters, the picture was reverse: these were significantly more common in peer-reviewed publications with a substantial amount of letters to the editor (case reports, peer-reviewed: 6.0%, 219/3635 vs pre-prints: 0.9%, 35/3805; P<0.001; letters, peer-reviewed: 17.4%, 632/3635 vs pre-prints: 0.5%, 19/3805; P<0.001). Overall, in both peer-reviewed publications and pre-prints the most addressed research areas were prevention and control (26.1%, 950/3635 vs 42.4%, 1615/3805; P<0.001) and diagnosis (21.5%, 781/3635 vs 25.3%, 962/3805).

Among pre-print publications, the 8.6% (329/3805) have been converted into peer-reviewed journals, up to the covered dates, among which the two study designs most published were the observational study (87.8%, n=289/329) and the RCT (5.6%, n=18/329) while the research areas of most interest were prevention and control (40.7%, n=134/329) and diagnosis (31.6%, n= 104/329).

### 2) Characteristics of COVID-19 ongoing research in the registers (trials and systematic reviews)

**Table 2** shows that, in the Clinicaltrials.gov, 1621 of the 339863 records were focused on COVID-19 disease (0.5%). In turn, 652 of the 32553 records included in the Chinese Clinical Trial Registry (ChiCTR), were focused on COVID-19 disease (2%). No other register had over 200 records of COVID-19 research.

**Table 2.**
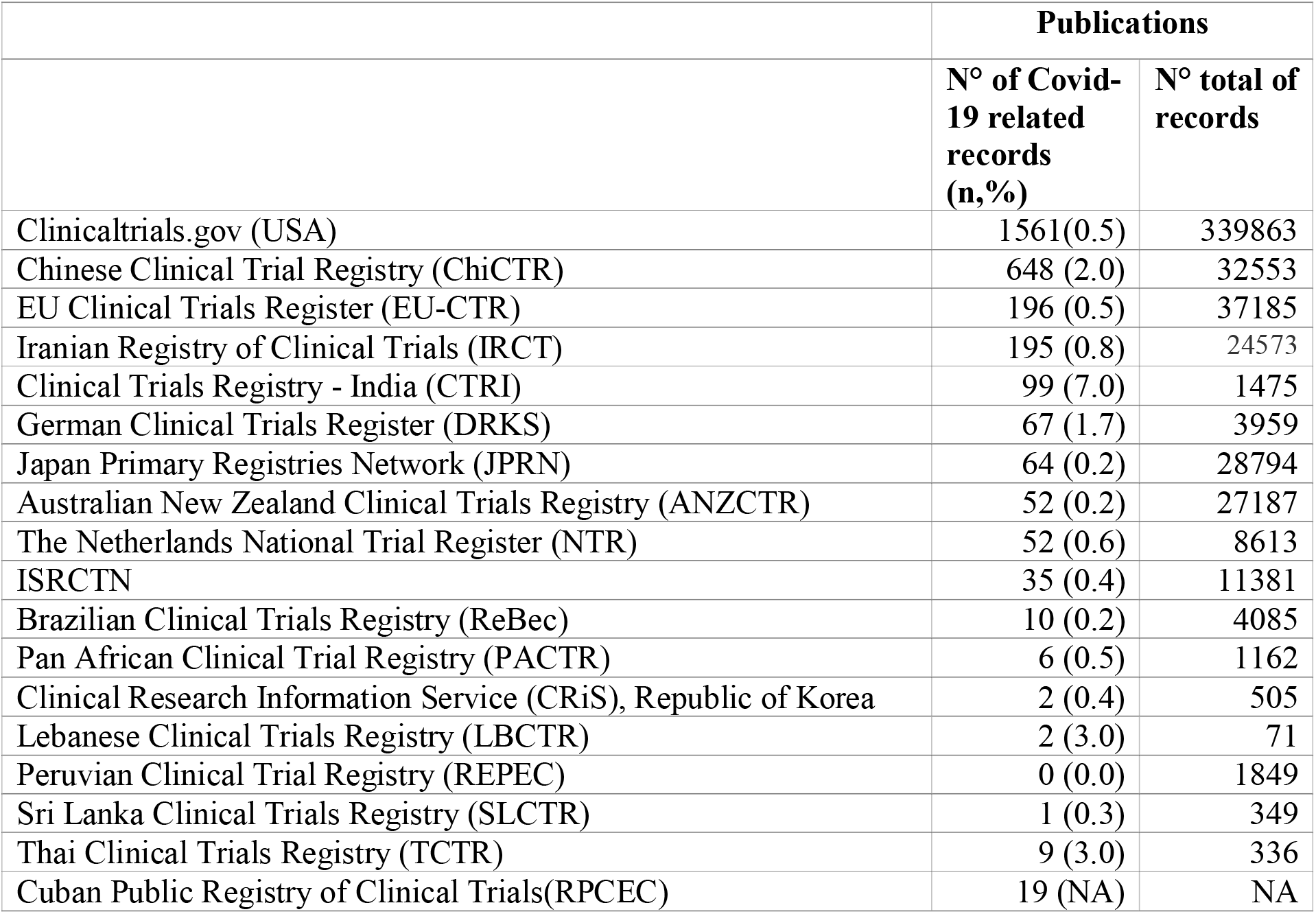
Primary Registries.

|  | **Publications** |  |
| --- | --- | --- |
|  | **N° of Covid-19 related records (n,%)** | **N° total of records** |
| Clinicaltrials.gov (USA) | 1561(0.5) | 339863 |
| Chinese Clinical Trial Registry (ChiCTR) | 648 (2.0) | 32553 |
| EU Clinical Trials Register (EU-CTR) | 196 (0.5) | 37185 |
| Iranian Registry of Clinical Trials (IRCT) | 195 (0.8) | 24573 |
| Clinical Trials Registry - India (CTRI) | 99 (7.0) | 1475 |
| German Clinical Trials Register (DRKS) | 67 (1.7) | 3959 |
| Japan Primary Registries Network (JPRN) | 64 (0.2) | 28794 |
| Australian New Zealand Clinical Trials Registry (ANZCTR) | 52 (0.2) | 27187 |
| The Netherlands National Trial Register (NTR) | 52 (0.6) | 8613 |
| ISRCTN | 35 (0.4) | 11381 |
| Brazilian Clinical Trials Registry (ReBec) | 10 (0.2) | 4085 |
| Pan African Clinical Trial Registry (PACTR) | 6 (0.5) | 1162 |
| Clinical Research Information Service (CRiS), Republic of Korea | 2 (0.4) | 505 |
| Lebanese Clinical Trials Registry (LBCTR) | 2 (3.0) | 71 |
| Peruvian Clinical Trial Registry (REPEC) | 0 (0.0) | 1849 |
| Sri Lanka Clinical Trials Registry (SLCTR) | 1 (0.3) | 349 |
| Thai Clinical Trials Registry (TCTR) | 9 (3.0) | 336 |
| Cuban Public Registry of Clinical Trials(RPCEC) | 19 (NA) | NA |

**Table 3** reported characteristics of the two primary registries with most volume of research COVID-19 related. The most investigated study type were the interventional studies in Clinicaltrials.gov as well as in ChiCTR (respectively, 60% and 50%), with the phase 3 most reported in Clinicaltrials.gov (15%). Regarding recruiting status, the most prevalent was “ongoing” across all registers (min-max: 47%-99%).

**Table 3.** Characteristics of Primary Registries with most volume of research COVID-19 related. (Absolute frequencies). Na*: records not available due to not possible search

| Characteristics |  | Clinicaltrials.gov<br>(n total=1621)<br>n, % | Chinese Clinical<br>Trial Registry<br>(ChiCTR)<br>( n total=652)<br>n, % | EU register<br>( n total=196)<br>n, % |
| --- | --- | --- | --- | --- |
| **Country** | Africa | 56 (3.5) | - | - |
|  | Central America | 3 (0.2) | - | - |
|  | East Asia | 120 (7.4) | - | - |
|  | *Japan* | 4 (0.2) | - | - |
|  | Europe | 628 (38.7) | - | 196 (100.0) |
|  | Middle east | 83 (5.1) | - | - |
|  | North America | 380 (23.4) | - | - |
|  | *Canada* | 44 (2.7) | - | - |
|  | *United States* | 326 (20.1) | - | - |
|  | *Mexico* | 18 (1.1) | - | - |
|  | North Asia | 13 (0.8) | - | - |
|  | Pacifica | 10 (0.6) | - | - |
|  | South America | 48 (2.9) | - | - |
|  | South Asia | 20 (1.2) | - | - |
|  | East Asia | 18 (1.1) | 652 (100.0) | - |
| **Study Type** | Interventional | 940 (57.9) | 323 (49.5) | - |
|  | Observational | 663 (40.9) | 260 (39.9) | - |
|  | Other (i.e.Patient Registries) | 125 (7.7) | 69 (10.6) | - |
| **Phase** | Early Phase 1 | 19 (1.2) | 218* (33.4) | - |
|  | Phase 1 | 91 (5.6) | 13 (2.0) | 6 (3.1) |
|  | Phase 2 | 375 (23.1) | 8** (1.2) | 99 (50.5) |
|  | Phase 3 | 241 (14.9) | 3*** (0.5) | 75 (38.3) |
|  | Phase 4 | 55 (3.4) | 67 (10.3) | 33 (16.8) |
|  | Not applicable | 282 (17.4) | 212 (32.5) | - |
| **Recruiting status** | Recruiting | 795 (49.0) | 308 (47.2) | 194 (98.9) |
|  | Complete | 76 (4.7) | 53 (8.1) | 0 |
|  | Suspended/temporarily halted | 7 (0.4) | 16 (2.5) | 0 |
|  | Other (e.g.. withdrawn) | 8 (0.5) | 275 (42.2) | 1 (0.5) |

Legend: Data were collected as reported from Primary registers.
\* phase 0 for ChiCTR
\*\*3 trials were I-II phase
\*\*\*1 trial was II-III phase

**Table 4** provided reporting characteristics of PROSPERO database about the registered COVID-19 related systematic reviews. Almost all are focused on humans (99.7%) with the two most research areas addressed to the treatments (19.1%) and health impacts (16.6%).

**Table 4.**
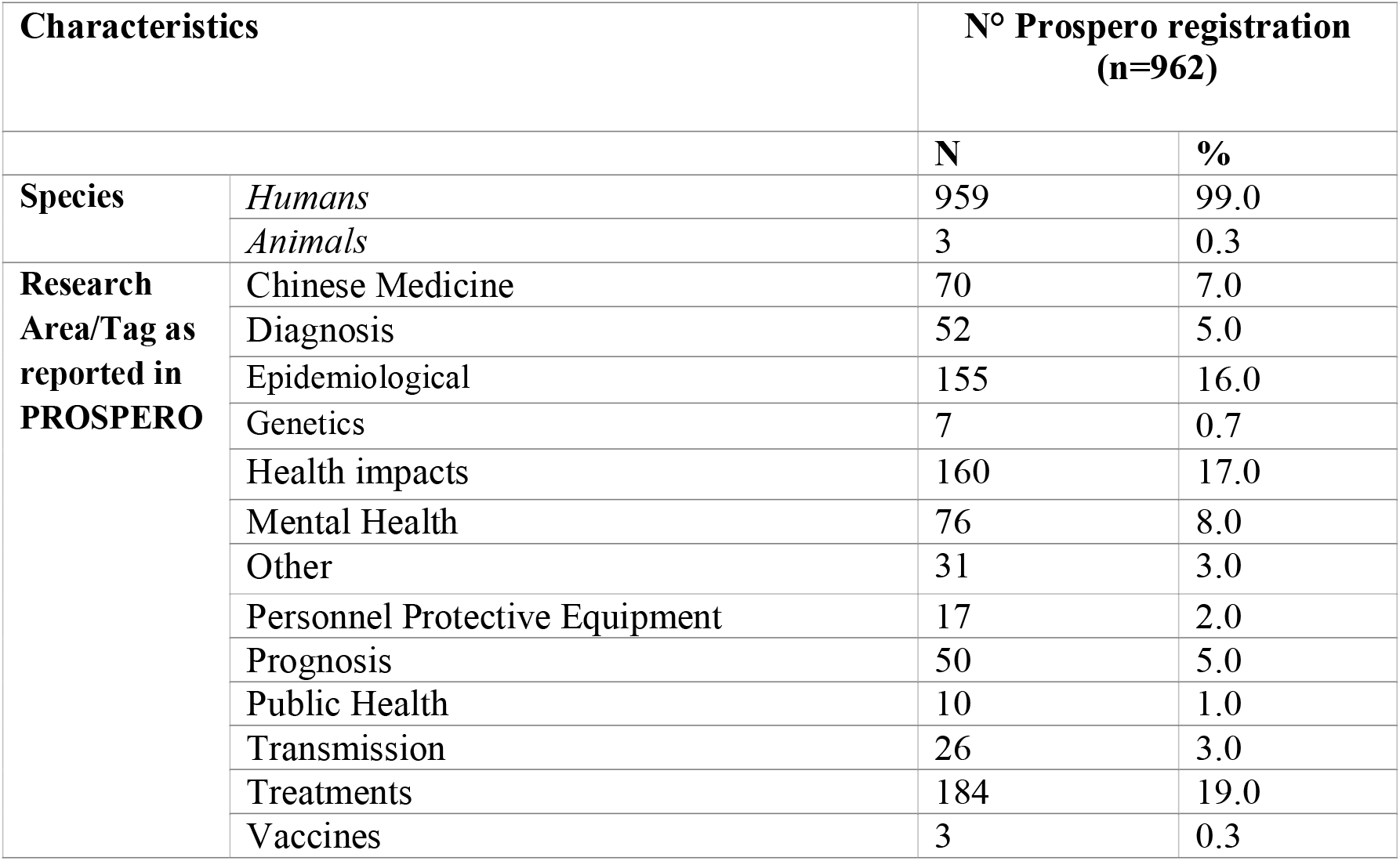
Characteristics of COVID-19 protocol of systematic reviews.

## Discussion

The spread of COVID-19 has rapidly been matched by an impressive rise in related publications in both peer-reviewed journals and pre-print servers (respectively, 3805 and 3635 records in about five months, with both a significant statistically increase [*p*< 0.001]). However, it is critical for clinicians, researchers and other stakeholders (e.g., government officers) to be able to identify, in a timely manner, accurate, reliable and trustworthy research. The snapshot we obtained reveals the publication and study register data on the first scientific endeavors to pandemic. To fill the void given by the absence of the published primary studies, immediate opinions (i.e., letters, editorials and comments) has virulently been published in the peer-reviewed literature, whereas empirical studies, including epidemiologic aiming to discover the foundations for biologic, prognostic and prevention and controls research areas, have been more commonly seen among pre-prints, up to May 20, 2020.

Our findings reflect a staged maturity of the research development. Even though the number of recorded publications was found high and increasingly sharply in just a few months (not counting the last few weeks likely due to indexation delays), we also found few higher-hierarchy evidence publications like systematic reviews/meta-analysis (n=29) and randomized controlled trials (n=8) in peer-reviewed journals. Others have found, similarly, a limited number of high-quality study designs on the COVID-19 disease (14, 24, 25). There has been insufficient time to design, approve, execute, and report such type of study designs; hence, it is probable that the percentage of these can rise over the upcoming months. For instance, the numbers of trials protocols that were found registered as “ongoing” in clinicaltrials.gov as well as in ChiCTR (respectively, n=795 and n=308) provide and indicator of that upward trend. From both registers, we retrieved a total of 2273 trials: the growth of this amount is remarkable since it has been reported on February 22, 2020, a total of 171 COVID-19-related interventional trials (138 from ChiCTR and 33 from ClinicalTrials.gov (26) and on March 24, 2020, a total of 614 COVID-19-related interventional trials (471 from ChiCTR and 143 from ClinicalTrials.gov) (27). This amount is justified by the breakthrough news from pandemic countries (such as China) which drew significant attention to drug interventions (such as azithromycin, hydroxychloroquine and antiviral drug)(28).

Besides, an important amount of systematic review protocols has been also registered in PROSPERO (n=962), and mainly focused on research treatments. However, these reviews can be limited by the low amount of clinical trials, or the lower quality of them (14). Indeed, secondary literature can be supported by indirect evidence represented by primary studies involving different patients from those of interest (i.e., COVID-19) as suggested by the Grading of Recommendations Assessment, Development and Evaluation (GRADE) approach (29).

The clinical development process to find answers, treatments and vaccines for pandemics is stronger if based on the highest levels of evidence such as systematic reviews and randomized controlled trials. Companies and universities have been doing their best to accelerate experimental drugs and vaccines for COVID-19 through their pipeline (30). The World Health Organization (WHO)’s Blueprint list recognized coronaviruses as disease of priority importance: given the public health emergencies of international concern and the absence of efficacious drugs and vaccines, coronaviruses disease are considered to need accelerated research and development (31). For the duration of the Coronavirus disease (COVID-19) outbreak several journals supported the immediate available publications through preprints posted online (32-34). However, until the pre-print is not published in a notable peer-reviewed journal it might get stuck in the limbo of credibility. We found only the 8% of pre-prints been published as peer-reviewed publications: does the explanation for such low percentage rely on too little time for convertion into a published version and a consequent delay of publication? Or is it merely a waste of research? On one hand, the time it takes for a completed manuscript to be published traditionally can be lengthy, up to 166 days for a preprint to be posted as DOI of a journal article (35). Article publication delay, often due to constraints associated with the peer review process, can prevent the timely dissemination of information in a global health emergency as COVID-19 pandemic (35). On the other hand, studies that remain unpublished, research’s “dark side of the moon”, could represent a waste of efforts and money annually invested in health and medical research globally (36, 37). It also happens that important results in some areas (such as mathematics) have been published only on preprint servers as ArXiv (38, 39). The potential harm from posting erroneous provisional research is one reason why the medical community was so cautious about preprints in the first place (40). However, to date, it is interesting to observe that pre-prints are actually gaining more attention and citations than the corresponding peer-reviewed article (41). For the COVID-19 related articles, it is still soon to analyze citation trends; yet, the publication trends here reviewed (i.e. up to May 20, 2020) showed a higher amount of preprints than peer-reviewed publications and higher percentage of empirical research among preprints. This contrasts with the findings from Lv et al up to February 6, 2020, such as those that articles in peer-reviewed journals accounted for 77.1% of the study publications, pre-prints 14.1%, while 8% of the articles were merely posted online (42). Explanations for these discrepancies can rely on the substantial amount of recent empirical studies that have been more rapidly available in preprint serves and not in peer-reviewed forms, at least yet. A better fruition of the trusted science can be enhanced by the better utilization of preprints. Preprints aim to give a new, faster, and iterative alternative or complement to the current journal publication and peer review system, proposing catalyze biomedical discovery, support career advancement, and improve scientific communication (35). If all publishers endorsed preprint posting for research, this would become an intrinsic preliminary step prior to the publication giving a clear signal to all scientists that preprints are integral part to scientific communication (18). Tracking the pre-print into the publication process can allow to control the reliability of research and distinct it from the final peer-reviewed publication. Also, it might give benefit for both individual authors and the scientific community. The former can immediately share their research and getting attention early, the latter can have direct access to the most updated research, give feedback to improve manuscripts, arise discussion leading to new ideas, follow-up studies, or collaborations with other research groups (43). This might be viewed as a reinforcement and an essential step toward a more open and transparent peer review process which is the cornerstone of our scientific activities and the full-proof system that ensures only quality research papers released into the scientific community (19). Commentaries, letters to editors and other forms of non-research publications were found essentially in peer-reviewed publication venues. These publication types seem less prone for a preprint form, or on the contrary benefit highly from certification by external peer-review in terms of not being essentially biased or personal perspectives. In turn, on empirical research, informed readers can more objectively appraise the research methods, the data presented, the validity of its conclusions, or limitations that are applicable, either in preprint or peer-reviewed versions. To answer the pandemic needs, the best challenge in this emergency would be increase the amount of completed and published research and translate it into practice.

### Limitations of the study

This study only covers journals and publications indexed in PubMed: a comprehensive but not exhaustive health research database. For pre-prints we used the COVID-19 section of these servers to investigate the preprint versions at the time of our data extraction. Our data collection period was a tightly defined window (December 2019-May 2020). However, since no filter nor topics are present for selecting pre-prints categories and the average time taken to display search results is not always optimal in the servers (44), a preferred hand screening was made. This partly threatens the validity of the comparisons of study types between peer-reviewed and preprint publications An illustrative example may be that of the epidemiologic studies: in pre-prints, we recorded different study designs as mentioned in the Methods section (such as Case-Control Studies, Cross-Sectional Studies but also extending to biologic and mathematical models), whereas in PubMed we involved the regular MeSH term that includes a clustered study designs (such as Case-Control Studies, Cross-Sectional Studies). Besides, epidemiological studies are indexed in PubMed not as a “publication type” like RCTs; this can affect the reliability, accuracy, and speed of the indexation of epidemiological studies in PubMed, e.g. compared to pre-print servers. This may help explain why many studies in detected PubMed had no classification for study type yet (i.e. the sum of the articles indexed for study types was clearly below the total number of papers identified). Yet, clear-cut publication types like RCTs or letters to editors may have had a faster a more reliable indexation. It could be reasonable to infer that many of the studies without indexation for a publication or study type could be other than RCT or commentaries/letters (e.g. epidemiologic, secondary data analysis, etc). Although indexation of COVID-19 research has been prioritized, delays may apply and these may have been visible in the data we obtained for the weeks of May, 2020.

When needed (i.e. when search filters or tags could not be reliably applied), manual coding was applied by one researcher and verified by another rather than by two independent researchers. This is an option typical of rapid review types, and a compromise needed to accelerate the results in an issue in which the timeliness of the results is key and a certain degree of uncertainty on the precision of the results. We did not adjusted characteristics for the country’s population size, for example by using World Bank’s data in order to facilitate any comparison on the amount of studies per country (https://data.worldbank.org/).

## Conclusion

There is an unprecedented increase of publications amount on COVID-19 since the disease emerged. There is a delay of accurate, reliable and trustworthy publication research. The vast majority of scientific literature in the first five months of this epidemic outbreak is based on perspectives (e.g. letters to the editor or commentaries) or reported (i.e. secondary) data rather than primary data. As well, a large number of registered trials and systematic review with very few results were published so far. The research focused on effective preventive/control and therapeutic strategies are still emerging reflecting the initial response by the research community. For rapid responses, science should promote preprints as a challenge and opportunity to give direct access to their research by submit them to journals that accept preprints for their policies. However, at a time when journal editors and invited reviewers are more solicited than ever, screening and reviewing preprints might benefit both the authors and the scientific community and can provide the general public with timely access to scientific discussion, helping to reinforce scientific credibility (33). Cautions should be taken regarding the phenomenon of “publish or perish”: the efforts and time should be devoted to develop significant and clinically relevant research agendas rather than investing more time to publish whatever researchers can get into print.

## Data Availability

Data and code used in this study are available on OSF

https://osf.io/vby7x

## Author contributions

Conceptualization: SG, GC, TJ;

Data curation, SG, GC;

Methodology: TJ, SG, GC;

Project Administration: SG, GC;

Formal analysis: GC, SG;

Investigation: SG, GC, SB;

Writing original draft: SG, GC;

Writing – Review & editing: SG, GC, TJ, SB.

## Data availability

Data and code used in this study are available on OSF https://osf.io/vby7x

## Abbreviation

COVID-19: Coronavirus disease 19
SARS-COV-2: Severe Acute Respiratory Syndrome coronavirus 2
PROSPERO: international prospective register of systematic reviews
MESH: Medical Subject Headings
SRs: systematic reviews
RCTs: randomized controlled trials

